# Knowledge, Attitude and Utilisation of Modern Contraceptives among Female Senior High School Students in the Kpando Municipality, Ghana

**DOI:** 10.1101/2023.10.01.23296407

**Authors:** Amanda Debuo Der, Elvis Enowbeyang Tarkang

## Abstract

**Background:** Adolescent fertility regulation and pregnancy prevention is one of the most important healthcare issues of the twenty-first century. Contraceptive can potentially improve the quality of the lives of adolescents and their economic welfare by preventing unwanted pregnancies. Volta Region records one of the highest prevalence of teenage pregnancy and adolescents aged 15-19 years are the least acceptors of contraceptives in the Region. This study determined the knowledge, attitude and utilisation of modern contraceptives among female senior high school students in the Kpando Municipality.

**Methods:** A descriptive cross-sectional design was employed in this study with a stratified sample of 270 participants. Data were collected using a self-administered questionnaire and analysed using Stata Version 16. Descriptive statistics such as frequencies and percentages were used to summarise the data. Logistic regression analyses were used to test the association between the dependent and independent variables at the 0.05 significance level at a 95% confidence interval.

**Results:** The participants had adequate knowledge of modern contraceptives (78.5%) and their major source of information was the school (59.7%) with the most commonly known contraceptive being the condom (86.3%). School of attendance (AOR= 0.29), religion (AOR= 0.25), marital status (AOR= 0.47) and programme of study were significantly associated with knowledge of modern contraceptives. The attitude of respondents was also favourable (63%) but the majority reported that modern contraceptives could lead to bareness in the future (59.3%). Only school of attendance was significantly associated with attitude (AOR= 2.04). However, current utilisation of modern contraceptives was low (24.2%) and the majority revealed their source of modern contraceptives to be the pharmacy (63.1%). Also, school of attendance (AOR= 2.36), age (AOR= 2.45), marital status (AOR= 41.81) and grade level (AOR= 2.13) were significantly associated with the utilisation of modern contraceptives.

**Conclusion:** There was adequate knowledge and favourable attitude regarding contraceptives. However, utilisation was low. Health promotion campaigns to improve contraceptive use among SHS students in Kpando Municipality should target those in the lower grade levels, those between the ages of 12 and 17, those who are currently not in any relationship and include all students regardless of their programme of study.

## INTRODUCTION

Contraception is the use of any technique, drug, device or method to prevent unplanned pregnancy, unsafe abortion and sexually transmitted infections (STIs) such as chlamydia, gonorrhoea, syphilis and HIV/AIDS [1]. Improving the reproductive health of young women in developing countries necessitates access to safe and effective methods of fertility control; however, most rely on traditional rather than modern contraceptives; special attention is required to ensure that the contraceptive needs of vulnerable groups, such as unmarried young women, poor women, and rural women, are met, and that knowledge and access inequities are reduced [2, 3].

Contraceptive use has increased in many parts of the world, especially in Asia and Latin America, but continues to be low in Sub-Saharan Africa (SSA) and the use of modern contraception has risen from 48% in 1990 to 57% in 2012 worldwide [4]. Sub-Saharan Africa has the highest average fertility rate in the world but is recording a decline since the use of traditional methods of contraception has reduced [5]. However, failure to use contraceptives still contributes to an unacceptably high rate of undesired pregnancy among SSA adolescents with associated maternal and neonatal mortality/morbidity on the rise [6].

Adolescent fertility regulation and pregnancy prevention is one of the most important health-care issues of the twenty-first century as adolescent females aged 15-19 account for over 14 million births each year, 91% of these occur in low and middle-income countries and an estimated 3 million girls aged 15 to 19 undergo unsafe abortions every year [4, 7, 8]. Contraceptive use has the potential to improve the quality of the lives of adolescents and their economic welfare and the lack of contraceptive use and their unfamiliarity with contraception can lead to an increase in several unplanned pregnancies, abortions, STIs, societal and family rejection [7, 9].

Knowledge of contraception is universal in Ghana but usage is lowest among the youngest women aged 15-19 at 19% [10, 11]. The male condom is the most known modern contraceptive method and the most used [12, 13, 14, 15]. The proportion of family planning acceptors increased gradually from 18% in 2009 to 23% in 2014 in the Volta region with the age group 15-19 being the least acceptors [16]. The region became one of the two regions in Ghana with a high prevalence of teenage pregnancy in 2016 accounting for 15.5% of the national prevalence with the contraceptive acceptance among young people in the Kpando Municipality very low as compared to the adult population [17]. The HIV population among young adults 15-24 years increased between 2018 (34,219) to 2022 (40,497) and that of adolescents (10-19 years) stood at 21,439 in 2022 [18, 19]. The national prevalence of HIV reduced from 2.1% in 2018 to 1.8% in 2022 [18, 19]. The Volta region has 16,996 people living with HIV and the Kpando Municipality recorded a prevalence of 2.14% which is above the national median [19]. Against this backdrop, this study determined the knowledge, attitude and utilisation of modern contraceptives among female Senior High School (SHS) students in the Kpando Municipality in the Volta region of Ghana.

## METHODS

### Study site description

The total population stands at 58,552 in the Kpando Municipality and people between the ages of 0 and 19 make up 26,302 representing 44.92% [20]. The education service of the Kpando Municipality comprises two (2) Senior High Schools and one (1) technical school of which only two have female students. There are two (2) hospitals, five (5) health centres, seven (7) Community-based Health Planning and Services (CHPS) compounds, one (1) Reproductive Health Centre and two (2) maternity homes. All these health facilities provide some form of contraception and contraception counselling to the people.

### Study population

The study was carried out among females in SHSs in the Kpando Municipality. Only female students were used because they are vulnerable and suffer the consequences of unplanned pregnancy, unsafe abortions and STIs.

### Inclusion and exclusion criteria

#### Inclusion criteria

The study included female students who were studying in the Senior High Schools in Kpando Municipality and were available at the time of the study. Consent was sought from students who were 18 years and above and parental consent and child assent were sought from students who were below 18 years before the data were collected. Teachers, however, provided parental consent to students since they were in a boarding institution.

#### Exclusion criteria

Students who met the inclusion criteria but had medical challenges such as severe sickness or did not consent to be included were not part of the study.

### Study design

A quantitative study approach employing the descriptive cross-sectional study design was used. The purpose for choosing this study design was to ensure that, data for the study was collected at one point in time and used to measure the knowledge, attitude and utilisation of modern contraceptives among female senior high school students in the Kpando Municipality in 2020. This study design allows the findings to be generalized to a larger target population and is useful for the generation of hypothesis [21]. However, a change in knowledge, attitude and utilisation of modern contraceptives among female SHS students in the Kpando Municipality could not be measured. Adequate steps to deal with the weaknesses of the study design as a 5% non-response rate was added to the sample size and the purpose of the study was clearly explained to study participants before the data collection. Also, precautions including conflict of interest and selection of study participants among the student population were ensured to avoid bias.

### Sample size determination

The sample size for this study was calculated using the single proportion population formula by Cochran [22] 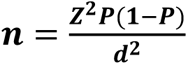

The sample size was estimated based on 21% (0.21) prevalence of contraceptive use among women of reproductive age in Ghana [23]. A confidence interval of 95%, a significance level of 5% (0.05 margin of error), a Z-score of 1.96 and a 5% non-response rate was used.

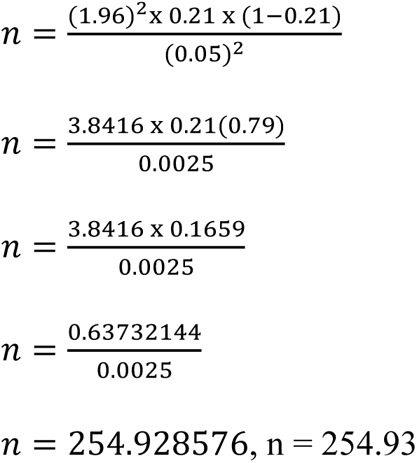

Adjusting for an anticipated 5% non-response rate, n = (254.93 x 0.05) + 254.93 = 12.7465 + 254.93 = 267.6765. A minimum total of 268 students was estimated, however, 270 students were recruited for the study.

### Sampling method

A multistage sampling technique, taking proportionate samples from the two SHSs in Kpando Municipal that had female students was used. The first stage involved purposively selecting the two schools that had female students. In the second stage, each class was treated as a stratum namely, SHS 1, SHS 2 and SHS 3. To ensure equal representation, proportionate samples were taken from each school and an equal number of female students were then selected from each class (the formula below was used to determine the number of female students to be selected from each school out of the sample size of 270). Female students who met the inclusion criteria were then selected from each stratum through simple random sampling. This was done by writing ‘Yes’ and ‘No’ on pieces of paper which were then squeezed and mixed and students who met the inclusion criteria were allowed to pick. Students who picked a ‘Yes’ were automatically part of the study.

Total number of female students from each school 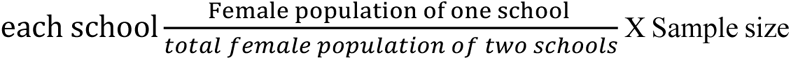 Female SHS 2 students in School B were not present at school during the data collection due to the double track system and hence the total female population of only SHS 1 and 3 was used for the calculation.

Therefore, the total number of female students from School B = 578/1290 ×268 = 120.08

This means that 120 female students were recruited in School B

Total number of female students from School A = 721/1290 ×268 = 149.79

This means that 150 female students were recruited in the School A

Number of female students from 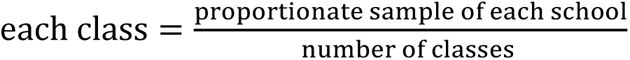

Number of female students from each class in the School B = 120/2 = 60 students from each class

Number of female students from each class in the School A = 150/3 = 50 students from each class

### Data collection procedure

A structured questionnaire was used to collect the data for the study on the knowledge, attitude and utilisation of modern contraceptives among the respondents. The questionnaire was self-administered and contained four sections. Section A contained 6 questions on the demographic characteristics of respondents while sections B, C and D contained 5 questions assessing respondents’ knowledge, 5 questions on attitude and 11 questions on utilisation of modern contraceptives respectively. Two females of the same educational level were trained to assist in the data collection. The questionnaire was adapted from similar studies [8, 11, 24, 25, 26]. It took participants about 20 minutes to complete the questionnaire. After completing the questionnaire, participants were appreciated by saying ‘thank you’. The questionnaires were pre-tested among 20 female students in a different SHS other than the study population to test the feasibility and reliability of the data collection instrument and all weaknesses of the instrument were improved before the actual data collection.

### Measures

The average score of each section was used as a guide in generating the scores. To generate the composite knowledge score, all ‘No’ and ‘Don’t Know’ responses were combined as ‘NO’. Nine (9) items were used to generate the score due to the presence of multiple-response questions and the average score was found to be 4. Therefore, respondents who scored below the average were considered to have had inadequate knowledge of modern contraceptives and those who scored the average and above were considered to have had adequate knowledge.

Further, five (5) questions were used to assess the attitude of respondents in this study and to get the composite attitude score, responses ‘Neutral’ and ‘Disagree’ to all questions were combined to be ‘Disagree’. The average score was found to be 1 and hence respondents who had zero (0) score were considered to have had an unfavourable attitude toward modern contraceptives and those who scored 1 and above were considered to have had a favourable attitude toward modern contraceptives. Also, on the current utilisation of modern contraceptives, one (1) item was used. Respondents who answered ‘yes’ were said to be currently using modern contraceptives and those who answered ‘no’ were considered not to be using modern contraceptives.

### Data analysis

Data collected were entered into Epi-data version 3.1 and exported into Stata version 16 for cleaning and analysis. Descriptive statistics such as frequencies and percentages were used to summarize the data. Logistic regression analysis was used to find the association between demographic variables and knowledge, attitude and contraceptive use. A test of probability value less than 5% (0.05) was considered statistically significant at a 95% confidence interval. Findings were presented in tables and graphs.

### Ethical issues

Ethical approval for this study was obtained from the University of Health and Allied Sciences Research Ethics Committee (UHAS-REC A.4[10] 19-20). Permission to carry out this study was also obtained from the Kpando Municipal Education Service and the administrative bodies of each school before the offset of the study. The content was disclosed and explained to participants in the language they best understood and their consent was obtained before commencement of the study. Participants 18 years and above signed an informed consent form and those below 18 signed a child assent form after their guardians had signed a parental consent form before they were allowed to participate in the study. Participation was voluntary and students could withdraw from the study at any time they wished. There were minimal risks associated with participating in this study (psychological risk due to the sensitive nature of the subject). There were also no direct benefits, however, findings will influence policies to the benefit of all female students in the Kpando Municipality. Names and details of participants were not linked to the data analysis and the findings to ensure anonymity. Furthermore, data management, storage, analysis and reporting were done using codes so as not to expose the details of the participants to ensure anonymity. Confidentiality was ensured by not disclosing information about participants to anybody.

## Results

### Demographic characteristics of the respondents

A total of 270 female students in SHSs in the Kpando Municipality took part in the study. Out of this, 150 (55.6%) were from the School A. The majority of the respondents 192 (71.1%) were between the ages of 12 and 17 years. Christians also represented the majority of the respondents 251 (93.0%). Among the respondents, 213 (78.9%) were single and the majority were Ewes 215 (79.6%). Most of the respondents were in SHS one (1) and SHS three (3) both representing 40.7% each with 124 (46.0%) studying home economics as shown in Table 1.

**Table 1:** Demographic characteristics of the respondents (n=270)

| Variable | Frequency (percentage) N (%) |
| --- | --- |
| **School** |  |
| School A | 150 (55.6) |
| School B | 120 (44.4) |
| **Age (Years)** |  |
| 12-17 | 192 (71.1) |
| 18-22 | 78 (28.9) |
| **Religion** |  |
| Christianity | 251 (93.0) |
| Islam | 19 (7.0) |
| **Marital status** |  |
| Single | 213 (78.9) |
| In intimate relationship | 57 (21.1) |
| **Ethnic group** |  |
| Ewe | 215 (79.6) |
| Others | 55 (20.4) |
| **Class (Grade level)** |  |
| SHS 1 | 110 (40.7) |
| SHS 2 | 50 (18.6) |
| SHS 3 | 110 (40.7) |
| **Program of study** |  |
| General art | 77 (28.5) |
| Technical skills | 43 (15.9) |
| Home economics | 124 (46.0) |
| Others | 26 (9.6) |

SHS: Senior High School

### Knowledge of modern contraceptives

Of the total respondents, the majority 263 (97.4%) had heard of some form of modern contraceptives before with condoms being the most known 227 (86.3%) and the intrauterine device (IUD) being the least known 29 (11.0%). Of the respondents, the majority, 157 (59.7%) first heard of modern contraceptives in school. Among the respondents who had heard of some form of modern contraceptives, the majority, 225 (85.6%) knew that modern contraceptives could help prevent unwanted pregnancy. However, 172 (65.4%) knew that modern contraceptives, especially condoms could help prevent both pregnancy and STIs (Table 2).

**Table 2:** Knowledge of modern contraceptives (n=270)

| Variable | Frequency (percentage) N(%) |
| --- | --- |
| **Heard of modern contraceptives before** |  |
| Yes | 263 (97.4) |
| No | 7 (2.6) |
| **If yes, which of the following have you heard before?** |  |
| **Pill** |  |
| Yes | 144 (54.8) |
| No | 119 (45.2) |
| **Condoms** |  |
| Yes | 227 (86.3) |
| No | 36 (13.7) |
| **Contraceptive injections** |  |
| Yes | 103 (39.2) |
| No | 160 (60.8) |
| **Contraceptive implants** |  |
| Yes | 45 (17.1) |
| No | 218 (82.9) |
| **Intrauterine device (IUD)** |  |
| Yes | 29 (11.0) |
| No | 234 (89.0) |
| **Emergency contraceptive pill** |  |
| Yes | 35 (13.3) |
| No | 228 (86.7) |
| **The first source of information** |  |
| School | 157 (59.7) |
| Parents | 10 (3.8) |
| Television | 53 (20.2) |
| Radio | 4 (1.5) |
| Friends | 23 (8.7) |
| Health facilities | 16 (6.1) |
| **Modern contraceptives help prevent unwanted pregnancy** |  |
| Yes | 225 (85.6) |
| No | 13 (4.9) |
| Don't know | 25 (9.5) |
| **Modern contraceptives especially condoms help prevent both pregnancy and STIs** |  |
| Yes | 172 (65.4) |
| No | 40 (15.2) |
| Don't know | 51 (19.4) |

On the overall knowledge of modern contraceptives, the majority of respondents 212 (78.5%) had adequate knowledge as shown in Figure 1.

**Figure 1:**
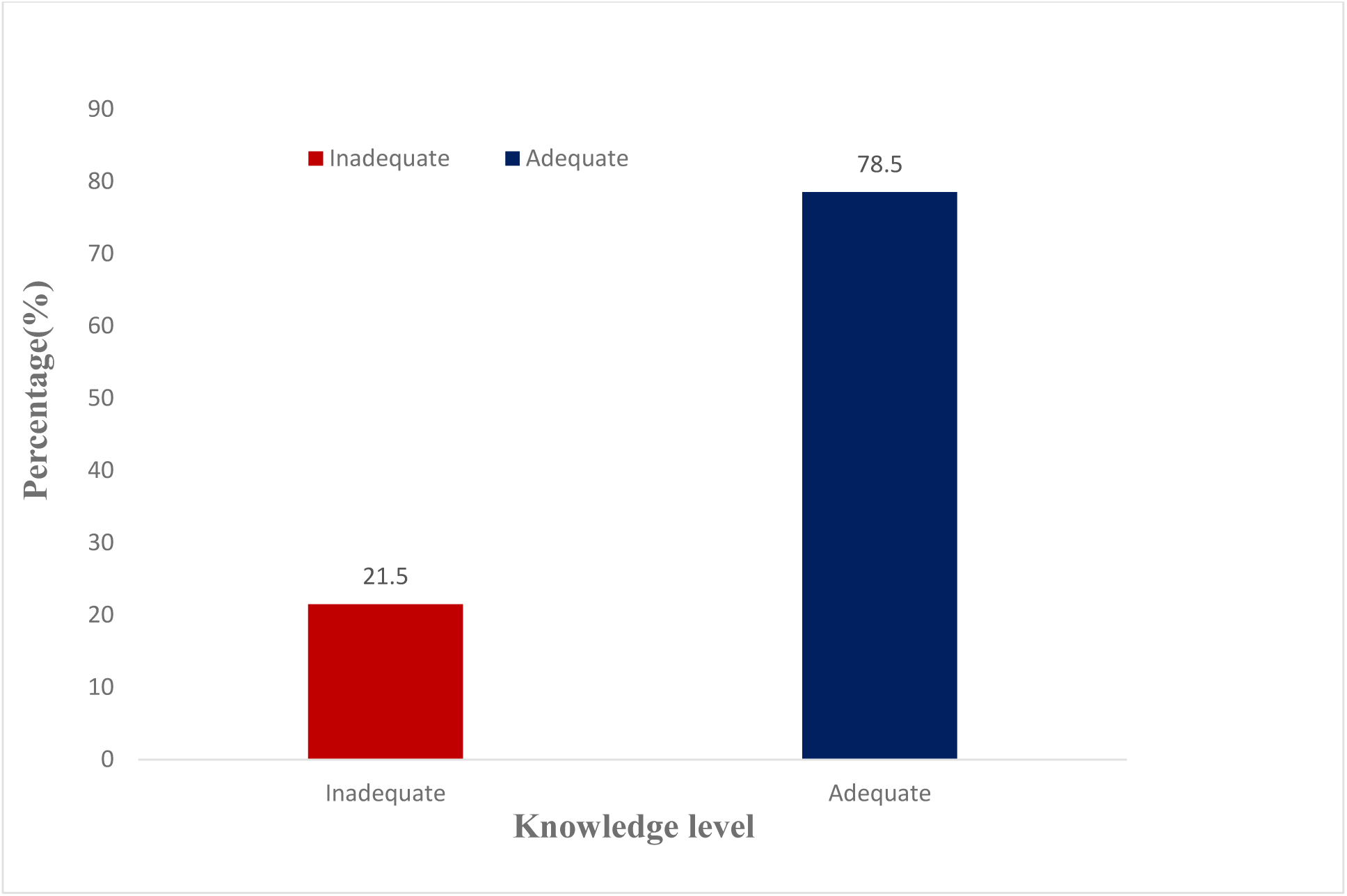
Overall knowledge of modern contraceptives.

### Association between demographic characteristics and knowledge of modern contraceptives

Table 3 shows the association between demographic characteristics and knowledge of modern contraceptives among the respondents. Respondents from School B were 71% less likely to have adequate knowledge [AOR=0.29(C.I: 0.16, 0.55); p< 0.001]; Muslims were also 75% less likely to have adequate knowledge than Christians [AOR=0.25 (C.I: 0.08, 0.76); p= 0.015]. Further, respondents who were in a relationship were 53% less likely to have adequate knowledge of modern contraceptives than their single counterparts [AOR=0.47 (C.I: 0.24, 0.91); p= 0.025]. More so, respondents who were studying technical skills and home economics were 73% and 70% less likely to have adequate knowledge of modern contraceptives [AOR=0.27(C.I: 0.10, 0.74); p= 0.011] and [AOR=0.30(C.I: 0.13, 0.70); p= 0.006] respectively.

**Table 3:**
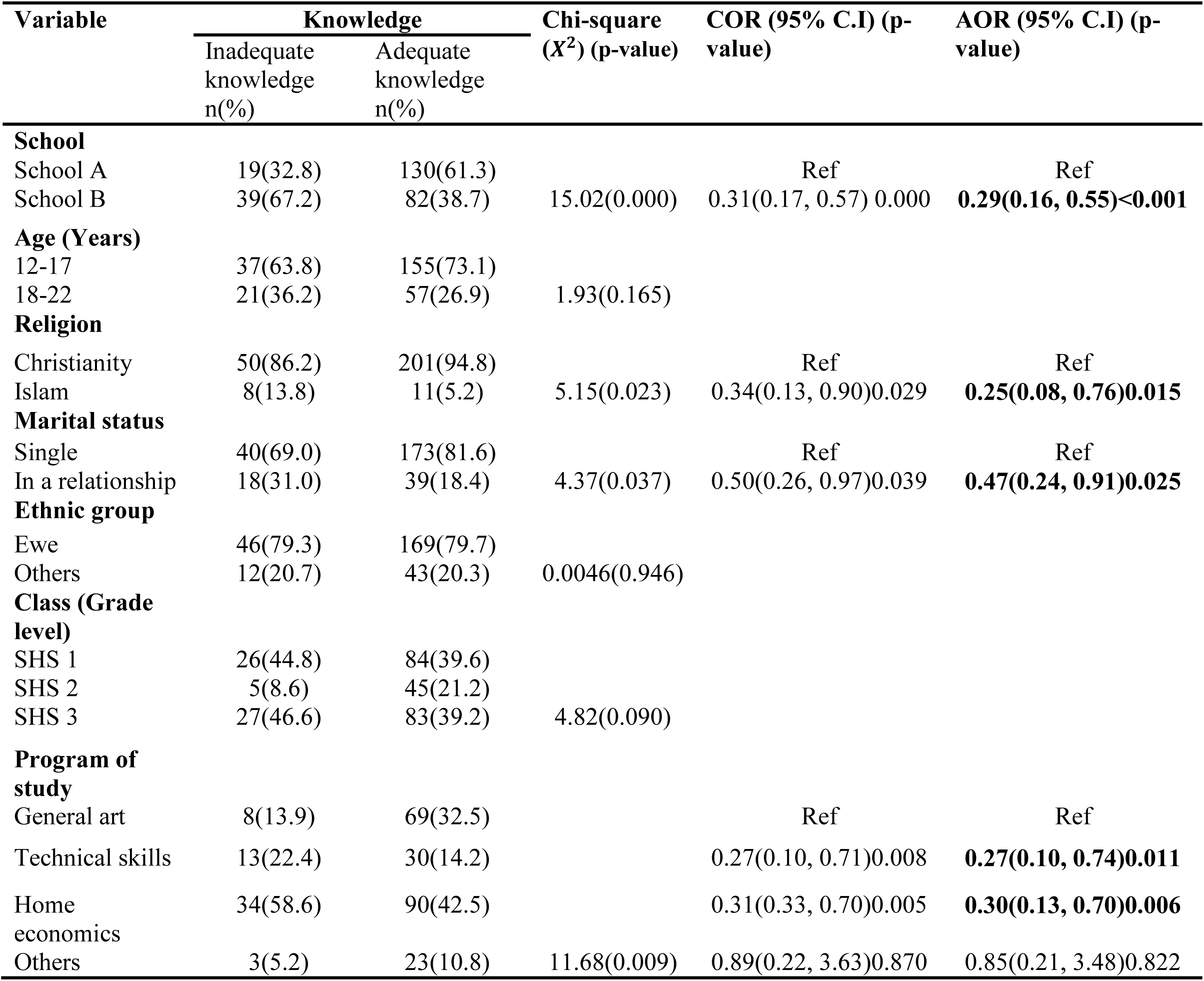
Association between demographic characteristics and knowledge of modern contraceptives.

### Attitude toward modern contraceptives

For description, responses, strongly disagree and disagree, and strongly agree and agree have been combined into disagree and agree respectively. Out of the 270 respondents, 84 (31.1%) agreed that modern contraceptives are expensive and the majority, 160 (59.3%) agreed that the use of modern contraceptives can lead to bareness in the future. In the same vein, the majority,177 (65.6%) agreed that they can approach a healthcare provider to get a modern contraceptive and 110 (40.7%) agreed to the assertion that people who use modern contraceptives are promiscuous while 114 (42.2%) agreed that modern contraceptives, especially condoms make sexual intercourse less pleasurable (Table 4).

**Table 4:** Attitude towards modern contraceptives (n=270)

| Variable | Frequency<br>(Percentage)<br>N (%) |
| --- | --- |
| **Modern contraceptives are expensive** |  |
| Disagree | 90 (33.3) |
| Neutral | 96 (35.6) |
| Agree | 84 (31.1) |
| **Modern contraceptive use leads to bareness in the future** |  |
| Disagree | 56 (20.4) |
| Neutral | 54 (20.0) |
| Agree | 160 (59.3) |
| **Cannot approach a health care provider for modern contraceptives** |  |
| Disagree | 54 (20.0) |
| Neutral | 39 (14.4) |
| Agree | 177 (65.6) |
| **People who use modern contraceptives are promiscuous** |  |
| Disagree | 56 (20.7) |
| Neutral | 104 (38.6) |
| Agree | 110 (40.7) |
| **Modern contraceptives especially condoms make sexual intercourse less pleasurable** |  |
| Disagree | 45 (16.7) |
| Neutral | 111 (41.1) |
| Agree | 114 (42.2) |

From Figure 2, the majority 170 (63%) of respondents had a favourable attitude toward modern contraceptives.

**Figure 2:**
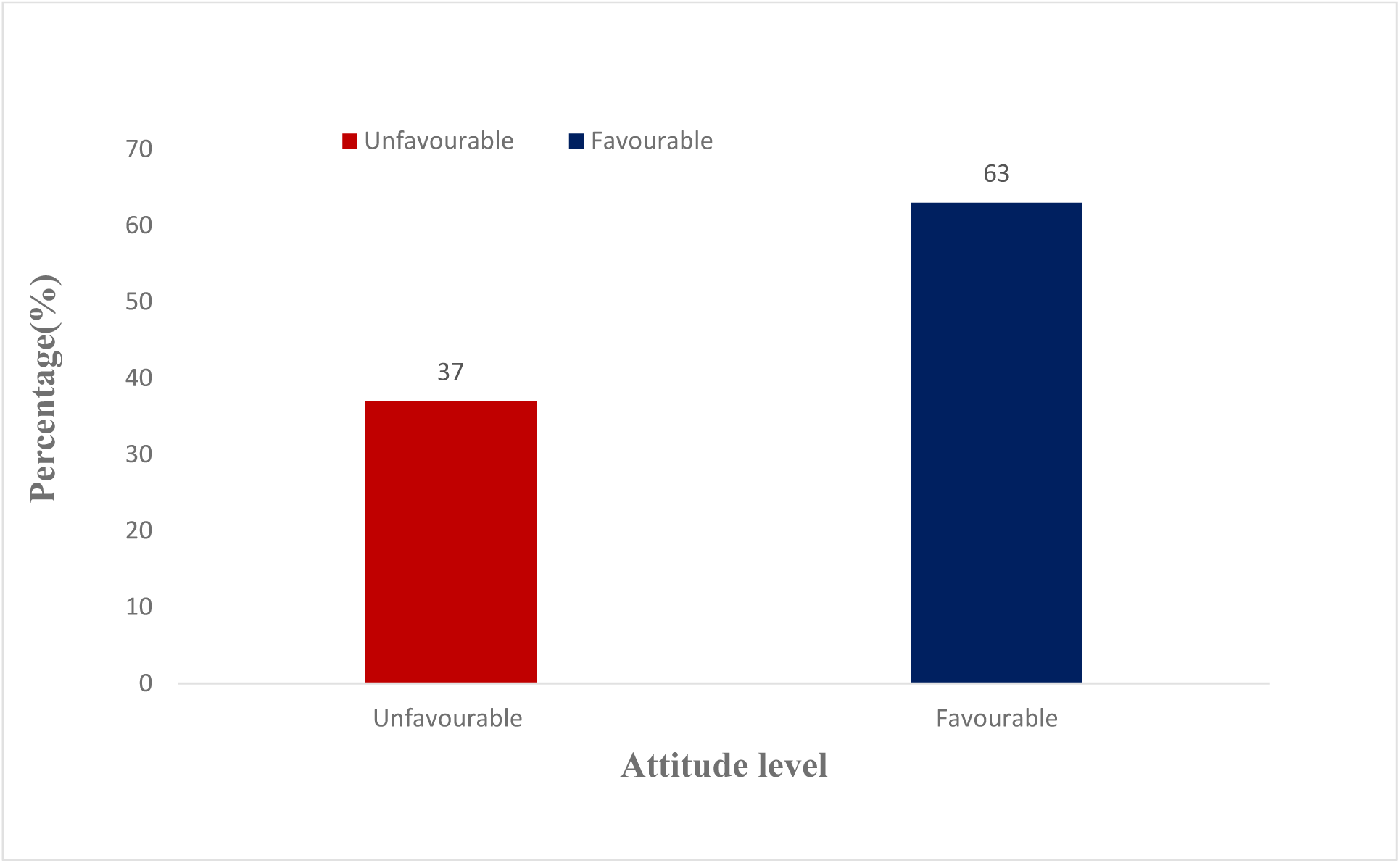
Overall attitude of respondents toward modern contraceptives.

### Association between demographic characteristics and attitude toward modern contraceptives

The association between demographic characteristics and attitudes toward modern contraceptives is shown in Table 5. Respondents from School B were 2 times more likely to have a favourable attitude toward modern contraceptives than those from School A [AOR=2.04 (C.I: 1.20, 3.48); p= 0.009].

**Table 5:** Association between demographic characteristics and attitude towards modern contraceptives.

| Variable | Attitude | | Chi-square<br>($X^2$) (p-value) | COR (95% C.I) (p-value) | AOR (95% C.I) (p-value) |
| --- | --- | --- | --- | --- | --- |
|  | Unfavourable attitude<br>n(%) | Favourable attitude<br>n(%) |  |  |  |
| **School** |  |  |  |  |  |
| School A | 67(67.0) | 82(48.2) |  | Ref | Ref |
| School B | 33(33.0) | 88(51.8) | 8.96(0.003) | 2.18(1.30, 3.64)0.003 | **2.04(1.20, 3.48)0.009** |
| **Age (Years)** |  |  |  |  |  |
| 12-17 | 71(71.0) | 121(71.2) |  |  |  |
| 18-22 | 29(29.0) | 49(28.8) | 0.00(0.975) |  |  |
| **Religion** |  |  |  |  |  |
| Christianity | 94(94.0) | 157(92.4) |  |  |  |
| Islam | 6(6.0) | 13(7.7) | 0.26(0.609) |  |  |
| **Marital status** |  |  |  |  |  |
| Single | 84(84.0) | 129(75.9) |  |  |  |
| In a relationship | 16(16.0) | 41(24.1) | 2.49(0.114) |  |  |
| **Ethnic group** |  |  |  |  |  |
| Ewe | 78(78.0) | 137(80.6) |  |  |  |
| Others | 22(22.0) | 33(19.4) | 0.26(0.610) |  |  |
| **Class (Grade level)** |  |  |  |  |  |
| SHS 1 | 32(32.0) | 78(45.9) |  |  |  |
| SHS 2 | 23(23.0) | 27(15.9) |  |  |  |
| SHS 3 | 45(45.0) | 65(38.2) | 5.41(0.067) |  |  |
| **Program of study** |  |  |  |  |  |
| General art | 32(32.0) | 45(26.4) |  | Ref | Ref |
| Technical skills | 8(8.0) | 35(20.6) |  | 3.11(1.28, 7.59)0.013 | 1.63(0.56, 4.74)0.367 |
| Home economics | 47(47.0) | 77(45.3) |  | 1.17(0.65, 2.08)0.606 | 0.70(0.33, 1.47)0.345 |
| Others | 13(13.0) | 13(7.7) | 8.85(0.031) | 0.71(0.29, 1.74)0.454 | 0.70(0.29, 1.73)0.054 |

### Utilisation of modern contraceptives

From Table 6, 82 (30.4%) of the respondents reported to have had sex before. Of these, 33 (40.2%) used a modern contraceptive during their first sex while the rest did not because the sex was either not planned (15.5%) or they could not afford a modern contraceptive (2.6%). In the same regard, 50 (61.0%) had used the condom before and only one person (1.2%) had used the IUD before with 12 (14.6%) admitting to having used none of the modern contraceptive methods before.

**Table 6:**
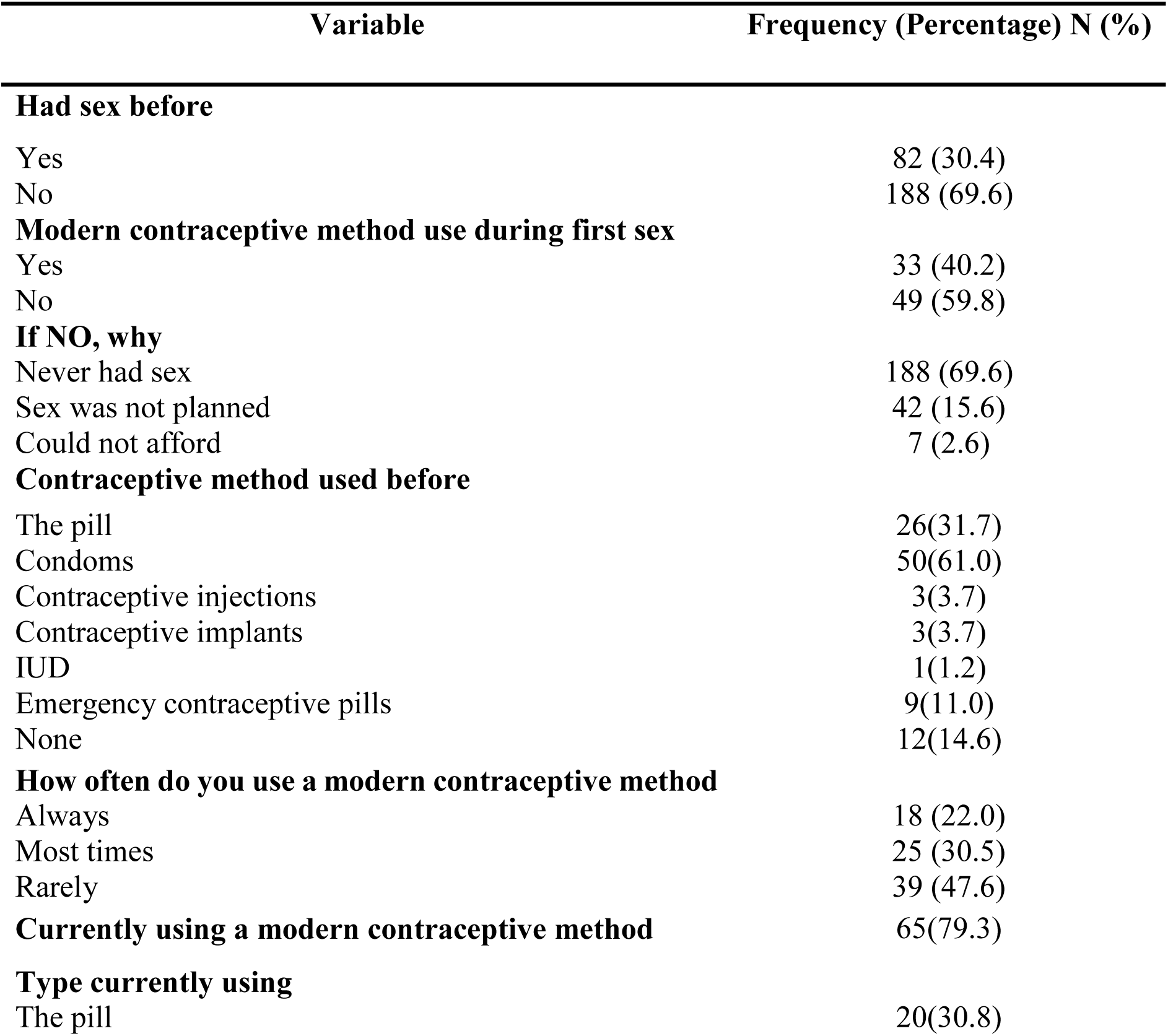
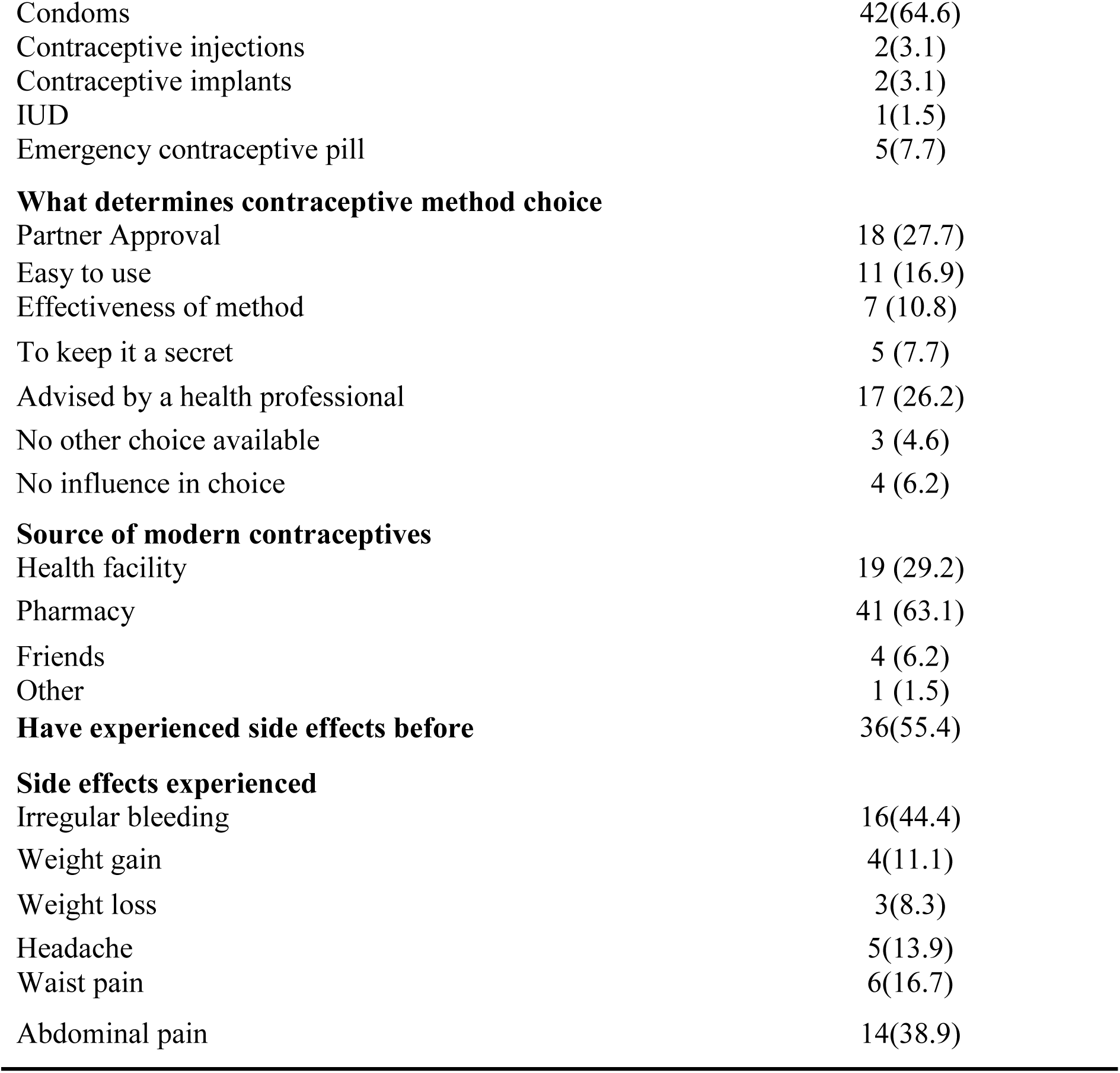
Utilisation of modern contraceptives (n=270)

| Variable | Frequency (Percentage) N (%) |
| --- | --- |
| **Had sex before** |  |
| Yes | 82 (30.4) |
| No | 188 (69.6) |
| **Modern contraceptive method use during first sex** |  |
| Yes | 33 (40.2) |
| No | 49 (59.8) |
| **If NO, why** |  |
| Never had sex | 188 (69.6) |
| Sex was not planned | 42 (15.6) |
| Could not afford | 7 (2.6) |
| **Contraceptive method used before** |  |
| The pill | 26(31.7) |
| Condoms | 50(61.0) |
| Contraceptive injections | 3(3.7) |
| Contraceptive implants | 3(3.7) |
| IUD | 1(1.2) |
| Emergency contraceptive pills | 9(11.0) |
| None | 12(14.6) |
| **How often do you use a modern contraceptive method** |  |
| Always | 18 (22.0) |
| Most times | 25 (30.5) |
| Rarely | 39 (47.6) |
| **Currently using a modern contraceptive method** | 65(79.3) |
| **Type currently using** |  |
| The pill | 20(30.8) |
| Condoms | 42(64.6) |
| Contraceptive injections | 2(3.1) |
| Contraceptive implants | 2(3.1) |
| IUD | 1(1.5) |
| Emergency contraceptive pill | 5(7.7) |
| **What determines contraceptive method choice** |  |
| Partner Approval | 18 (27.7) |
| Easy to use | 11 (16.9) |
| Effectiveness of method | 7 (10.8) |
| To keep it a secret | 5 (7.7) |
| Advised by a health professional | 17 (26.2) |
| No other choice available | 3 (4.6) |
| No influence in choice | 4 (6.2) |
| **Source of modern contraceptives** |  |
| Health facility | 19 (29.2) |
| Pharmacy | 41 (63.1) |
| Friends | 4 (6.2) |
| Other | 1 (1.5) |
| **Side effects experienced** |  |
| Irregular bleeding | 16(44.4) |
| Weight gain | 4(11.1) |
| Weight loss | 3(8.3) |
| Headache | 5(13.9) |
| Waist pain | 6(16.7) |
| Abdominal pain | 14(38.9) |

Moreover, 39 (47.6%) of the respondents said they rarely used modern contraceptive methods while 18 (22.0%) reported to have always used a modern contraceptive method during sexual intercourse. In addition, the majority 65 (79.3%) of respondents who have had sex before said they were currently using some form of modern contraception, with condoms being the most used method 42 (64.6%). The usage of these modern contraceptive methods was largely determined by partner’s approval 18 (27.7%) and least determined if no other choice was available 3 (4.6%), with most getting these modern contraceptives from the pharmacy 41 (63.1%). The majority, 51 (78.5%) reported that their partners agreed to their use of modern contraceptives. Of the 36 (55.4%) respondents who had experienced some side effects before, almost half, 16 (44.4%) reported irregular bleeding and the least, 3 (8.3%) reported weight loss as a side effect.

The utilisation of modern contraceptives was low, as the majority 205 (75.9%) of respondents were currently not using any form of contraceptive (Figure 3).

**Figure 3:**
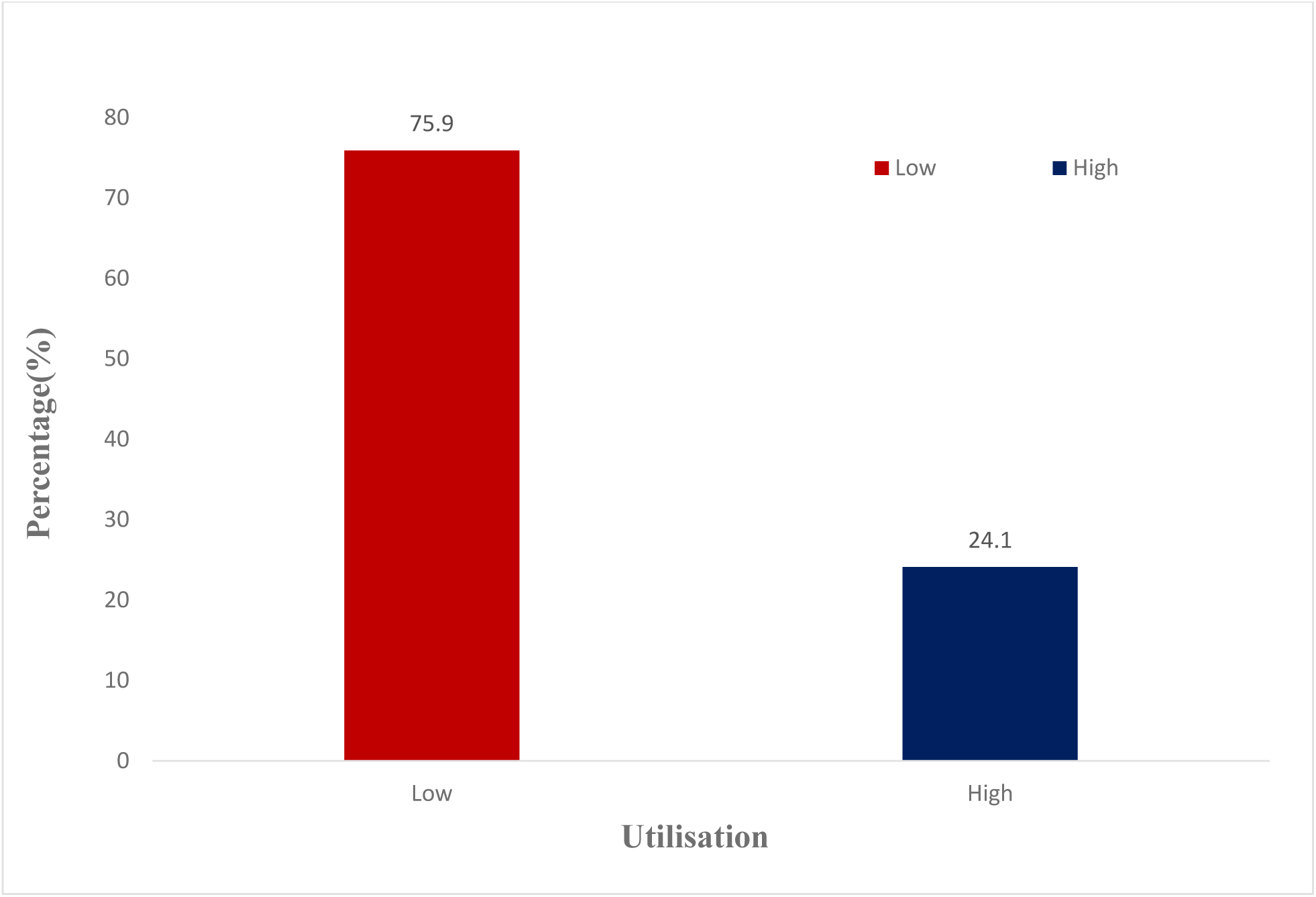
Utilisation of modern contraceptives.

### Association between demographic characteristics and utilisation of modern contraceptives

Respondents from School A were over 2 times more likely to use modern contraceptives [AOR= 2.36 (C.I: 1.32, 4.20); p= 0.004]. To add, respondents between the ages of 18 and 22 were 2 times more likely to use modern contraceptives [AOR=2.45(C.I: 1.33, 4.50); p= 0.004]. Also, respondents in a relationship were 42 times more likely to use a modern contraceptive method [AOR=41.81 (C.I: 18.58, 94.10); p< 0.001] and respondents in SHS 3 were 2 times more likely to use modern contraceptives [AOR=2.13 (C.I: 1.13, 4.01); p= 0.018] as indicated in Table 7.

**Table 7:**
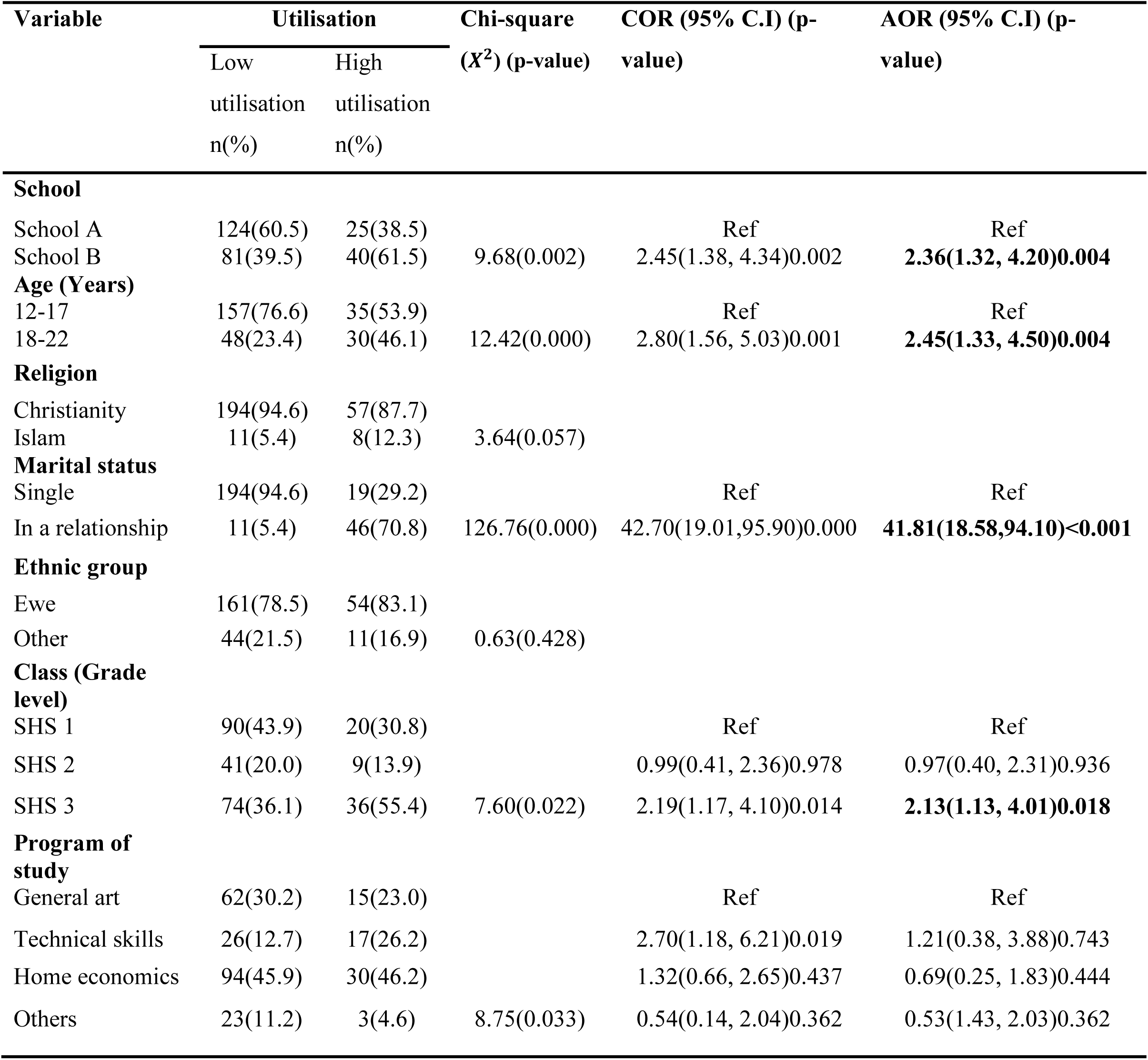
Association between demographic characteristics and utilisation of modern contraceptives.

## Discussion

### Knowledge of modern contraceptives

This study revealed the respondents to have adequate knowledge of modern contraceptives (78.5%). The majority (97.4%) had heard of one form of modern contraceptives before. The adequate knowledge in the current study is consistent with results from similar studies conducted in the Akuapem North Municipality of Ghana, in the Osun State of Nigeria and a nationally representative study in Ghana which reported similar levels of knowledge (96.1%, 61.5% and 99.8%) respectively [8, 11, 27]. The adequate knowledge in the current study could be attributed to the widespread campaigns on modern contraceptives that have influenced most people significantly especially female students as they wish to complete their education without an unplanned pregnancy or contracting an STI.

The most common modern contraceptive method known among respondents according to the current study was the condom (86.3%). This agrees with similar cross-sectional studies conducted in Northern Ghana among adolescents, Kintampo, Ghana and in the Limpopo province of South Africa, which reported that 74.8%, 84.0% and 58% of respondents respectively, knew of the condom [12, 13, 28]. The current finding could be attributed to the fact that campaigns on HIV/AIDS prevention have been focusing on condoms as it offer dual protection against unplanned pregnancy and STIs.

The major sources of information in the current study were school (59.7%), television (20.2%) and friends (8.7%). This corresponds with a cross-sectional study conducted in selected schools in the Osun state of Nigeria [8]. This could be due to the way some teachers factor in sex education when dealing with related subjects such as reproductive health in integrated science and biology. It could also be said that the media and friends have the potential to shape the behaviours of adolescents, concerning dating, sex and modern contraceptive use. Thus, the media could be used as a powerful tool necessary for the dissemination of information on modern contraceptives as it will reach a larger population. However, the current finding is in contrast with findings in the Limpopo province of South Africa among secondary school girls, which revealed parents to be the major source of information for respondents [28]. One could attribute this disparity with this current study to poor communication between parents and children and also due to the taboos surrounding the discussion of sex and contraceptives in some communities in Ghana and therefore children are compelled to learn about sexuality and contraception on their own through other means.

This study further revealed that the majority 172 (65.4%) of the respondents knew that modern contraceptives, especially condoms can help prevent both pregnancy and STIs. This is similar to a study conducted by Yeboah and Appai in the Akuapem North Municipality of Ghana [11]. Knowledge of the role of condoms is essential because translating it into contraceptive use behaviour will to a greater extent help to prevent STIs and unplanned pregnancies.

The school attended by respondents was found to be significantly associated with the knowledge of respondents. Those from School B were less likely to have adequate knowledge (AOR= 0.29, p< 0.001). This result is consistent with findings from a similar cross-sectional study conducted in the Osun state of Nigeria, which recorded differences in knowledge among selected schools [8]. This could be attributed to the different extracurricular activities such as school clubs in the two schools. School B clubs may possibly not focus on educating students on modern contraceptives. It is however necessary to incorporate the sexual health of students into all related extracurricular activities to provide the right information on sex and contraceptives to enable students to make informed choices to prevent unplanned pregnancies, unsafe abortions and STIs.

Another important finding from this study is that marital status was significantly associated with the knowledge of modern contraceptives as those who were in a relationship were 53% less likely to have adequate knowledge of modern contraceptives (AOR= 0.47, p= 0.025), and this is similar to that of a study conducted in Northern Ghana [13]. This finding is a grave public health concern as it could lead to an increase in unplanned pregnancies and STIs since those in a relationship are most likely to engage in sexual activities.

Further, religion was statistically associated with the level of knowledge of respondents as Muslim students were 75% less likely to have adequate knowledge (AOR= 0.25, p= 0.015). This corresponds with a study conducted in Kintampo, Ghana [12]. This finding could be due to the repressive nature of some religions regarding sex and contraceptive education especially among students, the majority of whom are not married. This, however, is a threat as young people are always curious about their developing bodies and want to explore their sexuality.

In addition, the program of study of the respondents was statistically significant with their knowledge level. Respondents studying technical skills and home economics were 73% and 70% less likely to have adequate knowledge (ARO= 0.27, p= 0.011) and (AOR= 0.30, p= 0.006) respectively. This is consistent with findings from a similar study conducted in Wuhan, China [29]. This could be attributed to the contents of these two programs as they are largely practical and focused and hence might not give respondents exposure to modern contraceptive information. This is, however, a concern as their inadequate knowledge level of modern contraceptives could lead them to make wrong choices regarding their sexual life which may lead to unplanned pregnancies, unsafe abortions and the possible contraction of STIs. It is, therefore, important for educational institutions to incorporate modern contraceptive education for all students regardless of their program of study.

### Attitude toward the use of modern contraceptives

According to this cross-sectional study, 63% of respondents had a favourable attitude toward modern contraceptives. This finding agrees with the results of a community-based cross-sectional study in Northwest Ethiopia among women of reproductive age, which reported that 58.8% of the respondents had a favourable attitude toward modern contraceptives [30].

Also, from this current study, 59.3% of respondents believed the use of modern contraceptives would lead to bareness in the future. This is in contrast with similar studies conducted in Northwest Ethiopia and in the Central region of Ghana as they both recorded low percentages of respondents who believed modern contraceptives could expose them to infertility in the future (24.5% and 18% respectively) [25, 30]. Although the participants in the current study had a favourable attitude towards modern contraceptives, the myth of being barren in the future could be a result of inadequate education on the safety of modern contraceptives, which could make them rely on traditional methods. This has the tendency to lead to a surge in cases of unwanted pregnancies, coupled with unsafe abortion and its complications as the efficacy of traditional contraceptive methods are not usually known. Consequently, campaigns on modern contraceptives need to spell out the associated myths and intensify programmes to render education that would demystify all misconceptions about modern contraceptives, emphasizing that unmarried people can safely use them.

Some respondents from the current study also agreed that modern contraceptives are expensive (31.1%) and this disagrees with a similar cross-sectional study conducted among SHS students in some selected schools in the Central region of Ghana, which revealed that 13% of students believed that modern contraceptives were expensive [25]. The current finding could be a result of poorer economic conditions in the current study location. However, the belief that people who use modern contraceptives are promiscuous was similar for both studies (40.7% and 40% respectively).

The school of attendance was significantly associated with the attitude of respondents toward modern contraceptives. Students in School B were 2 times more likely to have a favourable attitude toward modern contraceptives (AOR= 2.04, p= 0.009). It is important to note that even though respondents in School B were less likely to have adequate knowledge, they had a favourable attitude toward modern contraceptives. This implies that these students are more likely to use modern contraceptives if they are made accessible to them.

### Utilisation of modern contraceptives

Though the knowledge of modern contraceptives was high, the utilisation was considerably poor as only 24.1% reported currently using a modern contraceptive method. This is similar to cross-sectional studies conducted among female adolescents in the Volta region of Ghana and a nationally representative study in Ghana (18.7% and 34% respectively) [27, 31]. The low utilisation of modern contraceptives is a concern as this could lead to unwanted pregnancies, unsafe abortions and STIs among sexually active students. Young women must be given appropriate health education, including education about the use of modern contraceptives along with the risks associated with non-use.

The current study further revealed that the majority of respondents got contraceptives from pharmacies, and condoms were the most commonly used method. This is consistent with a similar study conducted in the North-East region of Ghana [15]. This could be because condoms are easily accessible at the pharmacy and also because condoms are the most appropriate contraceptive method used among unmarried people given its dual capacity to prevent STIs and unplanned pregnancy. The fact that the respondents were more knowledgeable about condoms, as compared to the other methods, might contribute to the reasons why condoms were mostly used.

The experience of side effects from the use of modern contraceptives by some respondents was inevitable. It was found from the study that 55.4% of respondents had experienced side effects. This differs from a similar study conducted among much older students in a public University in Ghana which revealed that 20% of respondents had ever suffered unpleasant negative side effects [32]. The experience of side effects among respondents in the current study could be due to incorrect use of the methods, especially as the pill was the second most used modern contraceptive method and irregular bleeding was the most reported side effect. It is always best to seek modern contraceptives from health facilities; therefore, adolescent-friendly corners should be situated in all health facilities to cater to the sexual needs of young people.

The school attended by respondents was found to be significantly associated with the utilisation of modern contraceptives. Students who were from School B were 2 times more likely to use modern contraceptives (AOR= 2.36, p= 0.004). It can be said that the use of modern contraceptives among students at School B could be attributed to their favourable attitudes toward modern contraceptives. This is impressive as it will in the long run lead to a reduction in unplanned pregnancies, unsafe abortions and STIs among students.

To add, the age of respondents was statistically significant with utilisation as those between the ages of 18 and 22 were more likely to use modern contraceptive methods (AOR= 2.45). This finding agrees with a study conducted in Ghana using the demographic and health survey data which found that older female adolescents were 3 times more likely to use modern contraceptives [3]. Perhaps, this is because older respondents are mature and enlightened than the younger ones in terms of the available contraceptive methods and the importance of contraceptive use. Also, older respondents are more likely to be sexually active and be in relationships than younger ones. This will help reduce the incidence of unplanned pregnancies, unsafe abortions and associated complications including STIs.

An important variable that was associated with modern contraceptive use according to this study was marital status, as respondents who were in a relationship were more likely to use modern contraceptives (AOR= 41.81) and this is consistent with a study conducted in Ghana using the demographic and health survey data [3]. This is understandable as respondents were students and would like to prevent unplanned pregnancies and STIs to enable them to focus on their education. Furthermore, class (grade level) was statistically significantly associated with the utilisation of modern contraceptives. Respondents from SHS 3 were 2 times more likely to use modern contraceptive methods (AOR= 2.13). This corroborates with findings from studies conducted in the Osun state of Nigeria and Ghana [8, 27, 33], which reported that students in higher classes were more likely to be sexually active and hence more likely to use modern contraceptives to prevent unplanned pregnancies, unsafe abortion and its complications and STIs.

## Conclusion

The knowledge of modern contraceptives was considerably adequate and the most commonly known were the condoms and the school attended by respondents, religion, marital status and programme of study were significantly associated with knowledge of modern contraceptives. Students had a favourable attitude toward modern contraceptives, however, most respondents believed that modern contraceptive use could lead to barrenness in the future. Only the school of respondents was significantly associated with attitude toward modern contraceptives. The utilisation of modern contraceptives was, however, low among the sexually active respondents and condoms were the most commonly used. The type and utilisation of modern contraceptives were largely influenced by partners and the main source of getting them was the pharmacy. The utilisation of modern contraceptives was significantly associated with the school of respondents, age, marital status and class (grade level).

## Recommendations

There is a need for healthcare workers to design campaigns that provide age-appropriate education on modern contraceptives. Also, sex education should be incorporated into the school curriculum to educate students and demystify the misconceptions surrounding modern contraceptives. In addition, adolescent-friendly corners should be established in hospitals to ensure that the right methods of modern contraceptives are provided to young people.

## Limitations of the study

The study addressed a sensitive topic where respondents were asked questions about their private sexual lives and this had the potential of introducing social desirability bias to the responses. The moral beliefs of society concerning sexuality, especially among young women could influence respondents’ ability to answer the questions in morally acceptable ways. However, confidentially was assured and participants were encouraged to individually respond to the questions and made aware that their responses were needed only for the research.

## Data Availability

All data produced in this present study are available upon reasonable request to the authors

## Acknowledgement

To the Ghana Education Service, Kpando Municipality and the students who participated in this study.

## References

1. Chebitok B. Knowledge of contraceptives, attitudes towards contraceptive use, and perceptions of sexual risk, among university students at a South African university. 2017. https://researchspace.ukzn.ac.za/handle/10413/15231.

2. Singh S, Darroch JE. Adding It Up: Costs and Benefits of Contraceptive Services— Estimates, New York. Guttmacher Institute and United Nations Population Fund (UNFPA) 2012. http://www.guttmacher.org/pubs/AIU-2012-estimates.pdf.

3. World Health Organization. Adolescent pregnancy. Fact sheet. 2014.

4. Nyarko SH. Prevalence and correlates of contraceptive use among female adolescents in Ghana. BMC Women’s Health. 2015;15(1):60.

5. Sharan M, Ahmed S, May J, Soucat A. Family Planning Trends in Sub-Saharan Africa: Progress, Prospects, and Lessons Learned. World Development Indicators. 2011:445– 69.

6. McCurdy RJ, Schnatz PF, Weinbaum PJ, Zhu J. Contraceptive use in adolescents in Sub-Saharan Africa: evidence from Demographic and Health Surveys. Conn Med. 2014;78(5).

7. Jahan U, Verma K, Gupta S, Gupta R, Mahour S, Kirti N, Verma P. Awareness, attitude and practice of family planning methods in a tertiary care hospital, Uttar Pradesh, India. Int J Reprod Contracept Obstet Gynecol. 2017;6(2):500–506. 10.18203/2320-1770.ijrcog20170370.

8. Tchokossa MA. Knowledge and Use of Contraceptives among Female Adolescents in Selected Senior Secondary Schools in Ife Central Local Government of Osun State. Int J Caring Sci. 2018;11(3):1647–61.

9. Appiah-Agyekum NN, Kayi EA. Students Perceptions of Contraceptives in University of Ghana. J Family Reprod Health. 2013;7(1):39–44.

10. Ghana Statistical Service and Ghana Health Service. Ghana Demographic and Health Survey. Accra, Ghana. 2015.

11. Yeboah T, Appai TP. Does knowledge of modern contraceptives and sexually transmitted infections affect contraceptive use and sexual behaviour? Evidence from senior high school girls in the Akuapem North Municipality, Ghana. GeoJournal. 2017;82(1):9–21.

12. Boamah EA, Asante KP, Mahama E, Manu G, Ayipah EK, Adeniji E, Owusu-Agyei S. Use of contraceptives among adolescents in Kintampo, Ghana: a cross-sectional study. Open Access J Contracept. 2014;5:7–15.

13. Yidana A, Ziblim S, Azongo TB, Abass YI. Socio-Cultural Determinants of Contraceptives Use Among Adolescents in Northern Ghana. Public Health Res. 2015;83–89. 10.5923/j.phr.20150504.01.

14. Kareem M, Samba A. Contraceptive use Among Female Adolescents in Korle-Gonno, Accra, Ghana. Gynecology & Obstetrics. 2016;6(12). 10.4172/2161-0932.1000414.

15. Dzantor EK, Serwaa A, Mahama A, Ayangba V, Agyeman YN, Kukeba MW, Adokiya MN. Contraceptive use among Students of a Health Training institution in the North-east Region of Ghana.

16. Himiede WW, Donne KA, Olayinka SI. Contraceptive Methods Accessed in Volta Region, Ghana, 2009 – 2014.

17. Wilson HW, Ameme DK, Ilesanmi OS. Contraceptive Methods Accessed in Volta Region, Ghana, 2009 – 2014. 10.1155/2017/7257042.

18. Ghana AIDS Commission. National and Sub-National HIV and AIDS Estimates and Projections. 2017.

19. Ghana AIDS Commission. Ghana’s HIV Fact Sheet 2022. National HIV Prevalence and Estimates. 2023.

20. Ghana Statistical Service. 2020 Population and Housing Census. General Report Volume 3B. 2021.

21. Omair A. Selecting the appropriate study design for your research: Descriptive study designs. J Health Spec. 2015;3(3):153.

22. Cochran WG. Sampling technique (3rd Edition). John Wiley and Sons. New York; 1977.

23. Benson P, Appiah R, Adomah-Afari A. Modern contraceptive use among reproductive-aged women in Ghana: prevalence, predictors, and policy implications. BMC Women’s Health. 2018;18(157):1–8. 10.1186/s12905-018-0649-2.

24. Cobbold JA. Factors influencing contraceptive use among adolescents in the Sunyani West district. 2018.

25. Hagan JE, Buxton C. Contraceptive knowledge, perceptions and use among adolescents in selected senior high schools in the central region of Ghana. J Sociol Res. 2012;3(2):170–180.

26. Grindlay K, Dako-Gyeke P, Ngo DT, Eva G, Gobah L, Reiger TS, Chandrasekaran S, Blanchard K. Contraceptive use and unintended pregnancy among young women and men in Accra, Ghana. 2018. 10.1371/journal.pone.0201663.

27. Oppong FB, Logo DD, Agbedra SY, Adomah AA, Amenyaglo S, Arhin-Wiredu K, Afari-Asiedu S, Ayuurebobi K. Determinants of contraceptive use among sexually active unmarried adolescent girls and young women aged 15–24 years in Ghana: a nationally representative cross-sectional study. BMJ Open. 2021 Feb 1;11(2):e043890.

28. Ramathuba D, Khoza LB. Knowledge, attitudes and practice of secondary school girls towards contraception in Limpopo Province. 2012. 10.4102/curationis.v35i1.45.

29. Zhang D, Bi Y, Maddock JE, Li S. Sexual and Reproductive Health Knowledge Among Female College Students in Wuhan, China. Asia-Pac J Public Health. 2010;22(1).

30. Kasa AS, Tarekegn M, Embiale N. Knowledge, attitude and practice towards family planning among reproductive age women in a resource limited settings of Northwest Ethiopia. BMC Res Notes. 2018;7–12. 10.1186/s13104-018-3689-7.

31. Akonor PY, Ayanore MA, Anaman-Torgbor JA, Tarkang EE. Psychosocial factors influencing contraceptive use among adolescent mothers in the Volta Region of Ghana: application of the Health Belief Model. Afr Health Sci. 2021;21(4):1849–1859.

32. Gbagbo FY, Nkrumah J. Family planning among undergraduate university students: a case study of a public university in Ghana. BMC Womens Health. 2019;19(1):12.

33. Kuenyefu PE. Utilization of Family Planning by Female Adolescents at Biakoye District in the Oti Region of Ghana [Masters Dissertation]. Ensign Global College; 2022.

